# A new method of Non-Ultrasound Monitoring of Ovarian Stimulation (NUMOS): Mission Possible! - a pilot study

**DOI:** 10.1101/2023.10.26.23297609

**Authors:** Iavor K. Vladimirov, Desislava Tacheva, Evan Gatev, Magdalena Rangelova, Martin Vladimirov

## Abstract

**Purpose:** This study aims to establish the viability of monitoring an appropriate and safe ovarian stimulation without the use of ultrasound and serum hormone testing.

**Method:** As a primary marker for monitoring of the ovarian response, we used urinary estrone-3-glucuronide (E1-3G) growth rate, which was self-measured by patients daily at home, with a portable analyzer, during the stimulation. For an adequate ovarian response, an average daily rate of increase of E1-3G was estimated to be within 25 - 77%. Ovulation trigger day was determined based on the length of the menstrual cycle.

The study included 24 women. Inclusion criteria were age < 41 years and AMH >1 ng/mL. A progestin-primed ovarian stimulation protocol (PPOS) with fixed doses of gonadotropins was used.

**Results:** The average female age was 32,9 years (±4.4), BMI 22,7 kg/m2 (±4,3), AMH 3,7 ng/ml (±2,6), stimulation days 10,6 (±1,1), collected oocytes 12,5 (±8,5), MII oocytes 10,6 (±7,8), fertilization rate 83,6% (±22,5), blastocyst 66,4% (±28,6), good quality blastocysts 31,6% (±16,9).

Absence of oocyte aspiration was found in one of the cases. There were no cases of OHSS and ovarian stimulation cancellation.

**Conclusion:** This is the first pilot study to successfully apply a new markers for ovarian stimulation monitoring.

## Introduction

Ovarian stimulation (OS) is defined as a pharmacological treatment with the intention of inducing the development of more follicles in the ovaries, and consequently obtaining multiple oocytes by follicular puncture (1). Control of ovarian stimulation (COS) is carried out through regular ultrasound examinations and blood sampling to monitor the dynamics of hormones. The main reasons for applying COS are to induce the maximum number of mature oocytes, thereby increasing the chance of live birth; to determine the appropriate time to apply the so-called ovulation trigger for the final maturation of the eggs and to prevent the risk of excessive ovarian response with ovarian hyperstimulation syndrome (OHSS), a rare iatrogenic complication of COS (2). Frequent OS monitoring visits are costly and time-consuming, as well as disrupting patients’ daily rhythm. In a study done in different European countries, 21 to 36% of all patients claim that they consider it hard to find time off work to implement fertility treatments. This is one of the main obstacle to seeking treatment (3).

To reduce the visits for ultrasound examinations, new methods are being developed to manage ovarian stimulation during in vitro treatment. They depend upon examining estrone-3-glucuronide levels in urine (4,5) and salivary hormone oestradiol and progesterone measurements (6).

Currently, two main stimulation approaches have been developed: Self-Operated Endovaginal Telemonitoring /SOET/ (7) and Controlled Ovarian Stimulation Monitoring by Self-Determination of Estrone-3-Glucuronide and Single Ultrasound (COSSESU). The implementation of these approaches is carried out with the active participation of patients and with the assistance of some elements of telemedicine (8). Some of the advantages that these two different approaches to ovarian stimulation provide, are the reduction of: costs of regular ultrasound and hormone tests; stress gained due to performing frequent blood tests in order to determine the values of different hormone levels in serum; time wasted in frequent clinic visits and traveling; direct non-medical expenses related to the use of a car, bus, train, hotel accommodation; food and risk of infection in situations similar to COVID-19.

The different approaches as described include elements of telemedicine. Even though many publications have been made in regards to using telemedicine in cardiology, dermatology, diabetes, and general practice, almost nothing has been reported regarding reproductive medicine. Interest has also increased tremendously due to the recent COVID-19 pandemic. We support the notion that telemedicine has a place in the treatment of infertility with the use of in vitro technologies (9).

This study aims to establish the viability of monitoring ovarian stimulation without the use of ultrasound and serum hormone testing so that the stimulation is appropriate and safe. We called that new method Non-Ultrasound Monitoring Ovarian Stimulation (NUMOS). Its main goal is to significantly reduce patient visits during stimulation and their respective costs.

## Materials and Methods

### Patient Selection Criteria

All patients (n = 24) underwent COH and IVF/ICSI procedures at the clinic (Sofia IVF clinic, SBALAGRM-SOFIA, Sofia, Bulgaria), during the period from November 2022 to January 2023. The study group of patients applied the Freeze-all policy. Their embryos were cryopreserved for later FET without fresh ET. All patients gave informed consent to participate in the program.

The study included 24 women with regular menstrual cycle (21-35 days) in 24 stimulated cycles. Inclusion criteria were age < 41 years and AMH >1 ng/mL. Exclusion criteria: patients with a severe male factor (spermatozoa <=1 million/ml), and endometriosis.

### Method

A progestin-primed ovarian stimulation protocol (PPOS) with fixed doses of gonadotropins was used. The starting doses were determined based on a preliminary assessment of the ovarian reserve, carried out in the previous 3 months.

Control of the adequacy of the ovarian response was based on monitoring the stimulation, as per the dynamics in the urine E3G ratio by regular determination of the E3G in the urine independently by the patients in home conditions. This was done with a small portable analyzer, which was easy to use and takes a short time to read E3G in the sample. The patient then sends the obtained result to the attending physician through a platform created for this purpose. A portable urine E3G analyzer and test strips were given to patients free of charge and patients received no compensation for participation. For an adequate ovarian response, an average daily rate of increase of E1-3G was estimated to be within 25 - 77% (10). In parallel, ultrasound examinations were performed, as the results of the study were not used to monitor the stimulation but were intended to minimize the risk to patients and the patient worries.

Criteria for determining the day for ovulation trigger and follicular puncture were based on the length of the menstrual cycle. It was calculated on the base of average length of the natural menstrual cycle for the three previous months, without the use of hormonal drugs for stimulation or contraceptives. We assumed that the middle of the menstrual cycle +/-1 day is the optimal day for follicular puncture.

Consider the following example of the new approach: with a 28-day menstrual cycle, the middle is the 14th day, respectively the ovulation trigger should be injected on the 12th day. If stimulation is started on day 2, then the number of stimulated days will be 10 days. If the stimulation is started from the 3rd day, then the number of stimulated days will be 9 days. In cycles longer than 30 days, stimulation lasts longer, with the day of follicular puncture being the 16th (+/-1) day of the menstrual cycle.

The deviation of one day was in accordance with the optimization of the work of the clinic related to the end of the week. The days of gonadotropin stimulation have no bearing on determining the day of follicular puncture.

A GnRH-agonist (Decapeptyl, Ferring) was used to trigger ovulation and was administered 34-36 hours before the follicular puncture (Fig 1).

**Fig. 1.**
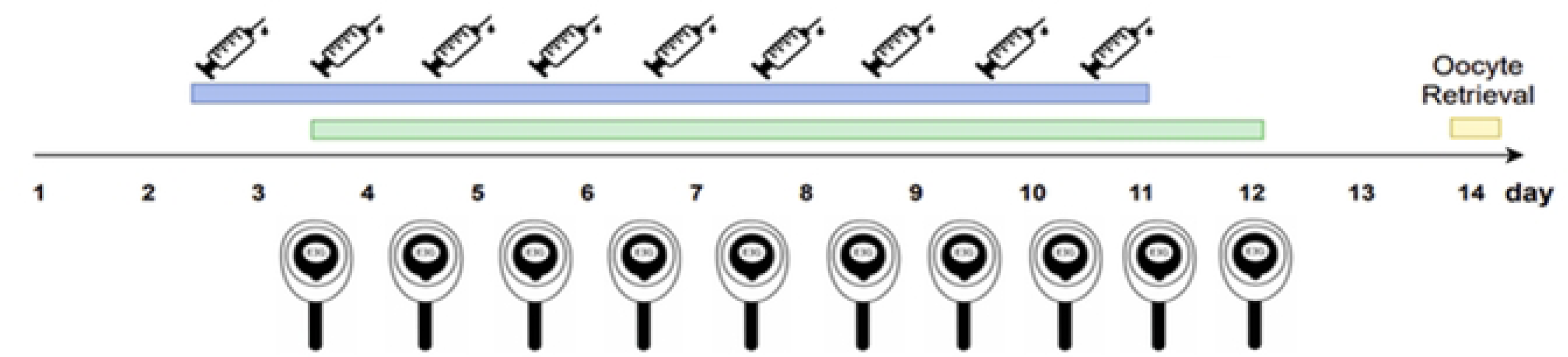
Non-Ultrasound Monitoring of Ovarian Stimulation (NUMOS)

Oocytes were fertilized by ICSI and all embryos were cultivated to blastocyst phase, and frozen.

### Statistical analysis

The ranges were determined based on quantile analysis.

## Results

The average female age was 32,9 years (±4.4), BMI 22,7 kg/m2 (±4,3), AMH 3,7 ng/ml (±2,6), stimulation days 10,6 (±1,1), collected oocytes 12,5 (±8,5), MII oocytes 10,6 (±7,8), fertilization rate 83,6% (±22,5), blastocyst formation 66,4% (±28,6), good quality blastocysts 31,6% (±16,9).

## Discussion

### Theoretical basis of the method

The success of the method is based on the application of criteria and methods that aim for the ovarian response to be: 1) appropriate to the applied stimulation doses, 2) effective, i.e. aspiration of oocytes sufficient in number and quality and 3) safe for the patients. The main factors on which the success of ovarian stimulation and its effectiveness depend are as follows:

### Choice of stimulation protocol

The application of PPOS makes treatment more convenient for patients by replacing some subcutaneous injections with oral medication.

The results of studies with different designs, different participant characteristics, and different progestins have established high-quality evidence supporting the effectiveness of progestins in obtaining similar numbers of oocytes with similar duration of stimulation and gonadotropin consumption compared to other stimulation protocols (11).

On the other hand, Guan et al. (12) in a meta-analysis of randomized controlled trials found that the number of oocytes retrieved, MII oocytes, and viable embryos was higher in PPOS protocols than in the control protocol. Based on a systematic review and meta-analysis, other authors found more obtained embryos in the PPOS protocol compared to standard stimulation protocols (13).

Next, the administration of progestins to inhibit ovulation with ovarian stimulation has been shown to be effective and safe with a low risk of OHSS. With the progestin protocol, the strategy of Freeze-all must be applied, in which case it is possible to apply a GnRH agonist as an ovulation trigger which significantly reduces the risk of OHSS. (14)

The American Society for Reproductive Medicine states that the risk of OHSS is about 1-2% and can be further reduced with the use of a GnRH agonist, triggering final oocyte maturation (15).

Ovulation triggering with a GnRH agonist almost completely eliminates the risk of OHSS without affecting oocyte and embryo quality and is therefore the trigger of choice. (16).

The above-mentioned advantages of the PPOS protocol and the possibility of administration of GnRH agonist as an ovulation trigger motivated our choice of the progestin protocol when applying the new approach to control ovarian stimulation.

### Determining the stimulation doses

Various algorithms have been developed to determine the starting doses of gonadotropins. The methods are based on preliminary determination of FSH and AMH levels, AFC, BMI, age, and history of smoking (17, 18, 19, 20,21). These algorithms for determining the initial stimulating doses aim for aspiration of 8-15 oocytes and eliminating the risk of OHSS in GnRH-agonist and antagonist protocols and performing the so-called fresh ET. In our new approach to ovarian stimulation control, we apply a PPOS protocol with mandatory embryo freezing. In this case, miscalculation of the stimulating doses and the use of higher ones did not affect the LBR in thawed ET, in contrast to fresh ET. On the other hand, when applying the Freeze-all strategy with a GnRH-agonist as an ovulation trigger, it would be desirable to aim for the retrieval of between 15 and 20 oocytes, which represents a good balance between safety and efficacy(22, 23).

### Monitoring the stimulation

Monitoring of the stimulation is based on the growth rate of E3G in the urine through its regular determination, independently by the patients in home conditions. The method is based on a relatively high correlation of estrone-3-glucuronide (E1-3G) urinary levels, which are a function of increasing estradiol levels during stimulation (24,25,26,27). The correlation was also established using the specific portable analyzer (4,10). Some components of the theoretical rationale developed by Vladimirov et al. (8) for monitoring ovarian stimulation by determining urinary E3G were applied. An appropriate response is identified with growth dynamics within established limits (28).

### Criteria for determining the day of follicular puncture

Our proposed new algorithm for determining the day follicular puncture is based on a thorough analysis of various scientific facts and regularities. We examined the relationship between the size of the follicles, the duration of the stimulation, and the length of the menstrual cycle, as well as the maturity of the oocytes, the quality of the embryos, and the success rate of the treatment.

A major method for predicting the possibility of aspiration of mature oocytes is the size of the follicles during ovarian stimulation. It is known that metaphase II (mature) oocytes can be aspirated from follicles of various sizes. In the literature, data on the ideal follicle size that determines the exact moment to apply the ovulation trigger are conflicting. No universally accepted follicle size threshold has been established to predict the presence of good competent oocytes. Overall, there is evidence that follicles with an average diameter between 16 and 22 mm are those most likely to contain MII oocytes, whereas small follicles rarely can generate MII oocytes. The frequency of mature oocytes increases with follicle size (329), however, in both relatively small and very large follicles, non-metaphase II oocytes can be aspirated. Mature oocytes are often aspirated from follicles larger than 16 mm in size. A greater number of immature oocytes are found in follicles smaller than 12 mm, and some authors recommend that such follicles should not be aspirated (30,31). Other authors found that mature oocytes can be aspirated from follicles with a smaller diameter, respectively between 11 to 14 mm (32,33,34).

Regarding the size of the follicles and the development of the embryos after fertilization, no relationship has been established because the results are contradictory (35,36). This suggests that the optimal follicle size may be population-specific.

In other studies, the authors found that in oocytes obtained from smaller follicles, the degree of polyspermy was higher after fertilization by conventional IVF, as well as a higher degree of fragmentation was observed, associated with a lower pregnancy rate (30). These effects may be due to reduced oocyte competence or due to safeguards against the entry of more than one spermatozoon and development during subsequent cleavage stages. Other authors did not confirm differences in oocyte maturation rate (35) or fertilization rate between small and large follicles. A mature oocyte can be obtained from a follicle measuring between 12 and 24 mm. Oocytes can be in metaphase II development, despite being derived from small follicles (0.3–0.9 mL volume), showing similar fertilization capacity and rate of blastocyst formation compared to oocytes of larger follicles (1-6 mL and >6 mL) (37). Some publications have demonstrated reduced fertilization rate and developmental competence of oocytes derived from very large (>23 mm) follicles, indicating the adverse effects of prolonged stimulation (38,39).

The facts presented above and our analysis give us reasons to believe that the size of the follicle from which a mature oocyte with normal characteristics can be aspirated varies widely. On the other hand, the main ultrasound criteria for determining an ovulation trigger, namely the presence of at least 3 follicles larger than 17 mm, also allows for a broader interpretation of the optimal day for follicular puncture.

### The question arises, does the day of follicular puncture depend on the duration of stimulation?

A number of authors have found that in the different protocols (long, antagonist or flare stimulation protocol), the duration of ovarian stimulation has no negative effect on the success rate in IVF/ICSI procedure (40). Optimal ovarian response and pregnancy outcome is associated with stimulation duration between 9-12 days (41).

Several authors have sought an answer to the question, whether changing the day for follicular puncture by one day from the optimal one (determined by ultrasound), has a negative effect on the quality of aspirated oocytes, respectively embryos and delivery rate. The purpose of the studies is to avoid performing follicular punctures on the weekend, and the results are related to determining the success rate based on fresh embryo transfer. Ben-Chetrit et al. (42) found that performing the follicular puncture a day earlier or later had no negative effect on the success rate using a GnRH-analogues protocol. Other authors have found that delaying hCG administration by 1 or 2 days does not worsen results in regard to the agonist protocol.

Another study has found that in patients with fixed days of stimulation in a long protocol, success rates were similar to those who had conventional ovarian stimulation monitoring (43). Similar results and conclusions also have been reported by authors applying an antagonist protocol in ovarian stimulation. Tremellen et al. (44) report that the early implementation of follicular puncture by 1 day from the optimum, leads to a decrease in the number of aspirated oocytes and created embryos. Delaying puncture by 1 day from optimal results in a non-significant increase in the number of oocytes collected and embryos created. However, the implementation of follicular puncture with the deviation of a day from the optimal one has no significant effect on the success rate based on live birth rates. The authors report that in a retrospective analysis of a large, randomized clinical trial (Engage) a one-day delay in HCG as an ovulation trigger does not negatively affect either the number of oocytes aspirated, or the pregnancy rate (45).

In support of the above studies (46,47,48,49) and the results of a meta-analysis of seven RCTs (50), the authors suggest that a delay in follicular puncture within 1-2 days when using a GnRH-antagonist protocol may result to aspiration of more mature oocytes without having an adverse effect on implantation and ongoing pregnancy rate. Mirzachi et al. (16) in their systematic review, reported that: moderate-quality evidence indicates that in freeze-all cycles, a moderate delay of 2–3 days in ovulation triggering may result in the retrieval of an increased number of mature oocytes without impairing the pregnancy rate. According to the authors, ovarian stimulation for frozen cycles is different in many aspects from conventional stimulation for fresh IVF cycles.

It is clear from the above that the duration of ovarian stimulation with gonadotropins can be within relatively wide limits, without having a negative effect on the quality of the oocytes, respectively pregnancy and live births.

For this reason, we looked for another approach and the use of other parameters to determine the ovulation trigger day and the follicular puncture day, respectively.

Already at the dawn of IVF technologies, a solution was sought through the so-called programmed oocyte retrieval to find a method that does not use US monitoring of ovarian stimulation. The authors used a predetermined day to apply an ovulation trigger without regard to the ovarian response. The authors report that this ovarian stimulation approach was effective in the treatment of infertility through the IVF procedure and was associated with significant practical and economic benefits (51, 52,53).

We hypothesized that the length of the menstrual cycle could be used to determine the duration of ovarian stimulation and that this would not negatively affect the success of the IVF procedure. It is known that the length of the menstrual cycle has a relationship with the aging of the ovary and its reserve (51). As ovarian function declines, the follicular phase is shortened, which is associated with lower AMH and AFC and poor IVF outcomes (16). On the other hand, menstrual cycle length has prognostic value for ovarian response in OS (54,55,56).

A study by Zhao et al. (57) remains in support of the thesis that the length of the menstrual cycle can be successfully applied as a marker to determine the day of follicular puncture. The authors determine the optimal day for triggering ovulation, based on determining the length of the follicular phase of the menstrual cycle. They define it as the difference between the length of the menstrual cycle and the length of the luteal phase, which is 14 days. 6,110 women with regular menstrual cycles were included in the study. From the analysis of the different groups, it can be seen that the days of stimulation vary between 8-12 days. A significant difference was found in the number of oocytes, fertilizations, embryos, and embryos of good quality in the different groups of patients, but this was not observed in comparisons based on pregnancies and births. The Q3 group, which has the best success rates, has an OS/FP (ovarian stimulation/follicular phase) ratio of 0.67-0.77. After analysis, it is evident that with a regular 28-day cycle, in the Q3 group, stimulation varying from 9 to 11 can be included, which are relatively wide limits.

Ours is the first pilot study to successfully apply a new approach to ovarian stimulation monitoring without the use of ultrasound and hormone analysis. This new method of monitoring and controlling ovarian stimulation based on other criteria such as the dynamics in the levels of E1-3G in urine and the length of the menstrual cycle. The in-depth analysis of the results of the studies, as well as our experience in using E1-3G in urine as a marker for the effectiveness of ovarian stimulation, motivated us to develop and successfully implement this new approach for monitoring the stimulation. The method can successfully replace the classical approach of monitoring stimulation through regular ultrasound examinations and analysis of serum hormone levels.

### Limitations

This study has some limitations. First, it is necessary to apply the method to a larger group of patients. Next, it is also necessary address the question, is this approach applicable only to the Freeze-all strategy or is it also possible to apply it with protocols that allow the implementation of fresh ET?

## Conclusion

This is a new approach for stimulation monitoring and does not exclude the use of standard methods. Its main goal is reducing patient visits during stimulation, respectively direct and indirect costs in IVF treatment.

In recent decades, we have witnessed the introduction of new technologies into assisted reproduction. The entry of artificial intelligence and telemedicine is already a fact. Established perceptions and approaches about how to control ovarian stimulation are constantly evolving. The development of new stimulation protocols and different strategies enables a look into a new era of IVF in the 21st century.

The new ovarian stimulation monitoring method developed and successfully implemented by us is part of this future. A two visits IVF treatment (TV/IVF) approach is now a reality, through which we will reduce the direct and indirect costs of infertility treatment. The current technology allows it!

## Data Availability Statement

The raw data supporting the conclusions of this article will be made available by the authors, without undue reservation.

### Ethics Statement

The studies involving human participants were reviewed and approved by the Ethics Committee of the SBALAGRM-SOFIA hospital. Informed consent for participation was written patients did not receive any compensation for participation.

## Author Contributions

IKV: Design, conception, analysis and interpretation of data, drafting of article, writing - review & editing, supervision. DT: Design, drafting of article, funding acquisition. MR: Acquisition of data, design. IG: Analysis and interpretation of data, writing - review & editing, visualization. MV: Conception, writing - review & editing, visualization. All authors approved the final manuscript. All authors contributed to the article and approved the submitted version.

## Funding

NA

## Conflict of Interest

The authors declare that the research was conducted in the absence of any commercial or financial relationships that could be construed as a potential conflict of interest.

## Acknowledgments

The authors would like to thank all the medical staff in the SBALAGRM-SOFIA for their assistance in data collection.

## Notes

### Competing Interest Statement

The authors have declared no competing interest.

### Funding Statement

The author(s) received no specific funding for this work.

### Author Declarations

The studies involving human participants were reviewed and approved by the Ethics Committee of the SBALAGRM-SOFIA hospital.

## References

1. Le KD, Vuong LN, Ho TM, Dang VQ, Pham TD, Pham CT, et al. A cost-effectiveness analysis of freeze only or fresh embryo transfer in IVF of non-PCOS women. Human Reproduction. 2018;33(10):1907–1914

2. Wu AK, Elliott P, Katz PP, Smith JF. Time costs of fertility care: The hidden hardship of building a family. Fertility and Sterility. 2013; 99:2025–2030. DOI: 10.1016/j.fertnstert.2013.01.145

3. Kelly J, Hughes CM, Harrison RF. The hidden costs of IVF. Irish Medical Journal. 2006; 99:142–143

4. Vladimirov I, Martin V, T Desislava T. Urine estrone–3-glucuronide (E3G) assay: Is there any place during ovarian stimulation for IVF cycles? Human Reproduction, Volume 36, Issue Supplement_1, deab130.669, 2021

5. Hart RJ, D’Hooghe T, Dancet EAF, Aurell R, Lunenfeld B, Orvieto R, et al. Self-monitoring of urinary hormones in combination with telemedicine - a timely review and opinion piece in medically assisted reproduction. Reproductive Sciences. 2021; 29:1–14. DOI: 10.1007/s43032-021-00754-5

6. Sakkas D, Howles CML, Atkinson L, Borini A, Bosch EA, Bryce C, et al. A multi-Centre international study of salivary hormone Oestradiol and progesterone measurements in ART monitoring. Reproductive Biomedicine Online. 2020; 42:421–428

7. Gerris J, Annick DA, Dhont N, Frank VF, Madoc B, Magaly BM, et al. Self-operated Endovaginal Telemonitoring versus traditional monitoring of ovarian stimulation in assisted reproduction: An RCT. Human Reproduction. 2014; 29:1941–1948. DOI: 10.1093/humrep/deu168

8. Vladimirov IK, Vladimirov M, Tacheva D. A new protocol for controlled ovarian stimulation monitoring by self-determination of Estrone-3-glucuronide and single ultrasound (COSSESU). Open journal of Obstetrics and Gynecology. 2021; 11:1217–1228

9. Berg WT, Goldstein M, Melnick AP, Rosenwaks Z. Clinical implications of telemedicine for providers and patients. Fertility and Sterility. 2020;114(6):1129–1134. DOI: 10.1016/j.fertnstert.2020.10.048.

10. Vladimirov I K, Tacheva D, Gatev E, Rangelov M and Vladimirov M. Viability of home monitoring of estrone-3-glucuronide (E1-3G) urine levels in controlled ovarian stimulation: A pilot study. Human Reproduction, Volume 38, Issue Supplement_1, June 2023, dead093.932, 10.1093/humrep/dead093.93230.

11. Ata B, Capuzzo M, Turkgeldi E, Yildiz S, La Marca A. Progestins for pituitary suppression during ovarian stimulation for ART: a comprehensive and systematic review including meta-analyses. Hum Reprod Update. 2021 Jan 4;27(1):48–66. doi: 10.1093/humupd/dmaa040.

12. Guan S, Feng Y, Huang Y and Huang J. Progestin-Primed Ovarian Stimulation Protocol for Patients in Assisted Reproductive Technology: A Meta-Analysis of Randomized Controlled Trials. Front Endocrinol (Lausanne). 2021 Aug 31;12: 702558. doi: 10.3389/fendo.2021.702558. eCollection 2021.

13. Cui L, Lin Y, Wang F, Chen C. Effectiveness of progesterone-primed ovarian stimulation in assisted reproductive technology: a systematic review and meta-analysis. Arch Gynecol Obstet. 2021 Mar;303(3):615–630. doi: 10.1007/s00404-020-05939-y. Epub 2021 Jan 12.

14. La Marca A and Capuzzo M. Use of progestins to inhibit spontaneous ovulation during ovarian stimulation: the beginning of a new era? Reprod Biomed Online. 2019 Aug;39(2):321–331. doi: 10.1016/j.rbmo.2019.03.212. Epub 2019 Mar 29.

15. Practice Committee of the American Society for Reproductive Medicine and Practice Committee of the Society for Assisted Reproductive Technology. Repetitive oocyte donation: a committee opinion. Fertil Steril 2014; 102:964–966.

16. Mizrachi Y, Horowitz E, Farhi J, Raziel A and Weissman A. Ovarian stimulation for freeze-all IVF cycles: a systematic review. Human Reproduction Update, Vol.26, No.1, pp. 119–136, 2020.

17. Popovic-Todorovic B, Loft A, Bredkjæer HE, Bangsbøll S, Nielsen IK, Andersen AN. A prospective randomized clinical trial comparing an individual dose of recombinant FSH based on predictive factors versus a ‘standard’ dose of 150 IU/day in ‘standard’ patients undergoing IVF/ICSI treatment. Human Reprod. 2003;18(11):2275–2282.

18. Popovic-Todorovic B, Loft A, Lindhard A, Bangsbøll S, Andersson AM, Andersen AN. A prospective study of predictive factors of ovarian response in ‘standard’ IVF/ICSI patients treated with recombinant FSH. A suggestion for a recombinant FSH dosage normogram. Human Reprod. 2003;18(4):781–787.

19. Howles CM, Saunders H, Alam V, Engrand P. Predictive factors and a corresponding treatment algorithm for controlled ovarian stimulation in patients treated with recombinant human follicle stimulating hormone (Follitropin alfa) during assisted reproduction technology (ART) procedures. An analysis of 1378 patients. Curr Med Res Opin. 2006;22(5):907–918.

20. Olivennes F, Howles CM, Borini A, et al. Individualizing FSH dose for assisted reproduction using a novel algorithm: the CONSORT study. Reprod Biomed Online. 2009;18(2):195–204.

21. Yovich JL, Alsbjerg B, Conceicao JL, Hinchliffe PM, Keane KN. PIVET rFSH dosing algorithms for individualized controlled ovarian stimulation enables optimized pregnancy productivity rates and avoidance of ovarian hyperstimulation syndrome. Drug Des Devel Ther. 2016 Aug 10;10: 2561–73. doi: 10.2147/DDDT.S104104. eCollection 2016.

22. Gerber RS, Fazzari M, Kappy M, Cohen A, Galperin S, Lieman H, et al.. Differential Impact of Controlled Ovarian Hyperstimulation on Live Birth Rate in Fresh Versus Frozen Embryo Transfer Cycles: A Society for Assisted Reproductive Technology Clinic Outcome System Study. Fertil Steril (2020) 114:1225–31.

23. Munch EM, Sparks AE, Zimmerman MB, Van Voorhis BJ, Duran EH. High FSH dosing is associated with reduced live birth rate in fresh but not subsequent frozen embryo transfers. Hum Reprod 2017;32: 1402–1409.

24. Rapi S, Fuzzi B, Mannelli M, Pratesi S, Criscuoli L, Pellegrini S, et al. Estrone 3-glucuronide chemiluminescence immunoassay (LIA) and 17beta estradiol radioimmunoassay (RIA) in the monitoring of superovulation for in vitro fertilization (IVF): correlation with follicular parameters and oocyte maturity. Acta Eur Fertil. 1992;23(2):63–8

25. Catalan R, Castellanos JM, Palomino T, Senti M, Antolin M, Galard RM. Correlation between plasma estradiol and estrone3-glucuronide in urine during the monitoring of ovarian induction therapy. Int J Fertil. 1989;34(4):271–5.

26. Alper MM, Halvorson L, Lasley B, Mortola J. Relationship between urinary estrone conjugates as measured by enzyme immunoassay and serum estradiol in women receiving gonadotropins for in vitro fertilization. J Assist Reprod Genet. 1994;11(8):405–8.

27. Gary S. Nakhuda, Ning Li, Zheng Yang, Sylvia Kang. At-home urine estrone-3-glucuronide quantification predicts oocyte retrieval outcomes comparably to serum estradiol. FS Rep 2023 Jan 28;4(1):43–48.

28. Simonetti S, Veeck LL, Jones HW Jr. Correlation of follicular fluid volume with oocyte morphology from follicles stimulated by human menopausal gonadotropin. Fertil Steril. 1985;44(2):177–80.

29. Bergh C, Broden H, Lundin K, Hamberger L. Comparison of fertilization, cleavage and pregnancy rates of oocytes from large and small follicles. Hum Reprod. 1998;13(7):1912–5.

30. Rosen MP, Shen S, Dobson AT, Rinaudo PF, McCulloch CE, Cedars MI. A quantitative assessment of follicle size on oocyte developmental competence. Fertil Steril. 2008;90(3):684–90.

31. Scott RT, Toner JP, Muasher SJ, Oehninger S, Robinson S, Rosenwaks Z. Follicle-stimulating hormone levels on cycle day 3 are predictive of in vitro fertilization outcome. Fertil Steril. 1989;51(4):651–4.

32. Mehri S, Levi Setti PE, Greco K, Sakkas D, Martinez G, Patrizio P. Correlation between follicular diameters and flushing versus no flushing on oocyte maturity, fertilization rate and embryo quality. J Assist Reprod Genet. 2014;31(1):73–7.

33. Akbariasbagh F, Lorzadeh N, Azmoodeh A, Ghaseminejad A, Mohamadpoor J, Kazemirad S. Association among diameter and volume of follicles, oocyte maturity, and competence in intracytoplasmic sperm injection cycles. Minerva Ginecol. 2015;67:397–403.

34. Salha O, Nugent D, Dada T, Kaufmann S, Levett S, Jenner L, Lui S, Sharma V. The relationship between follicular fluid aspirate volume and oocyte maturity in in-vitro fertilization cycles. Hum Reprod. 1998;13:1901–16.

35. Nogueira D, Friedler S, Schachter M, Raziel A, Ron-El R, Smitz J. Oocyte maturity and preimplantation development in relation to follicle diameter in gonadotropin-releasing hormone agonist or antagonist treatments. Fertil Steril. 2006;85:578–83.

36. Haines CJ, Emes AL. The relationship between follicle diameter, fertilization rate, and microscopic embryo quality. Fertil Steril. 1991;55:205–7.

37. Raine-Fenning N, Deb S, Jayaprakasan K, Clewes J, Hopkisson J, Campbell B. Timing of oocyte maturation and egg collection during controlled ovarian stimulation: a randomized controlled trial evaluating manual and automated measurements of follicle diameter. Fertil Steril. 2010;94:184–8.

38. Ectors FJ, Vanderzwalmen P, Van Hoeck J, Nijs M, Verhaegen G, Delvigne A, Schoysman R, Leroy F. Relationship of human follicular diameter with oocyte fertilization and development after in-vitro fertilization or intracytoplasmic sperm injection. Hum Reprod. 1997;12:2002–5.

39. Wertheimer A, Nagar R, Oron G, Meizner I, Fisch B, Ben-Haroush A. Fertility treatment outcomes after follicle tracking with standard 2-dimensional sonography versus 3-dimensional sonography-based automated volume count: prospective study. J Ultrasound Med. 2018;37(4):859–66.

40. Purandare N, Emerson G, Kirkham C, Harrity C, Walsh D, Mocanu E. The duration of gonadotropin stimulation does not alter the clinical pregnancy rate in IVF or ICSI cycles. Ir J Med Sci (2017) 186:653–657.

41. Sarkar P, Ying L, Plosker S, Mayer J, Ying Y, Imudia AN. Duration of ovarian stimulation is predictive of in-vitro fertilization outcomes. Minerva Ginecol. 2019 Dec;71(6):419–426. doi: 10.23736/S0026-4784.19.04455-1. Epub 2019 Nov 13.

42. Ben-Chetrit, A., Senoz, S., Greenblatt, E.M., 1997. In vitro fertilization programmed for weekday-only oocyte harvest: analysis of outcome based on actual retrieval day. J. Assist. Reprod. Genet. 14, 26– 31.

43. Nakagawa, K., Yamano, S., Senuma, M., Myogo, K., Yamazaki, J., Aono, T., 1997. Avoidance of oocyte retrieval on the weekend through the use of scheduled ovarian hyperstimulation for in vitro fertilization and embryo transfer. Fertil. Steril. 68, 787–79.

44. Tremellen, K.P., Lane, M., 2010. Avoidance of weekend oocyte retrievals during GnRH antagonist treatment by simple advancement or delay of hCG administration does not adversely affectIVF live birth outcomes. Hum. Reprod. 25, 1219–1224.

45. Levy MJ, Ledger W, Kolibianakis EM, Ijzerman-Boon PC, GordonIs K. it possible to reduce the incidence of weekend oocyte retrievals in GnRH antagonist protocols? Reprod Biomed Online. 2013 Jan;26(1):50–8. doi: 10.1016/j.rbmo.2012.09.014. Epub 2012 Sep 26.

46. Kolibianakis EM, Albano C, Camus M, Tournaye H, Van Steirteghem AC, Devroey P. Prolongation of the follicular phase in in vitro fertilization results in a lower ongoing pregnancy rate in cycles stimulated with recombinant follicle-stimulating hormone and gonadotropinreleasing hormone antagonists. Fertil Steril 2004;82:102–107.

47. Kolibianakis EM, Bourgain C, Papanikolaou EG, Camus M, Tournaye H, Van Steirteghem AC, Devroey P. Prolongation of follicular phase by delaying hCG administration results in a higher incidence of endometrial advancement on the day of oocyte retrieval in GnRH antagonist cycles. Hum Reprod 2005;20:2453–2456.

48. Morley L, Tang T, Yasmin E, Hamzeh R, Rutherford AJ, Balen AH. Timing of human chorionic gonadotrophin (hCG) hormone administration in IVF protocols using GnRH antagonists: a randomized controlled trial. Hum Fertil (Camb) 2012;15:134–139.

49. Levy MJ, Ledger W, Kolibianakis EM, Ijzerman-Boon PC, Gordon K. Is it possible to reduce the incidence of weekend oocyte retrievals in GnRH antagonist protocols? Reprod Biomed Online 2013;26:50–58.

50. Chen Y, Zhang Y, Hu M, Liu X, Qi H. Timing of human chorionic gonadotropin (hCG) hormone administration in IVF/ICSI protocols using GnRH agonist or antagonists: a systematic review and metaanalysis. Gynecol Endocrinol 2014;30:431–437.

51. Vassena R, Vidal R, Coll O, Vernaeve V. Menstrual Cycle Length in Reproductive Age Women Is an Indicator of Oocyte Quality and a Candidate Marker of Ovarian Reserve. Eur J Obstet Gynecol Reprod Biol (2014) 177:130–4. doi: 10.1016/j.ejogrb.2014.03.027

52 Braude P R, Bright M V, Douglas C P, Milton P J, Robinson R E, Williamson J G, Hutchison J. A regimen for obtaining mature human oocytes from donors for research into human fertilization in vitro. Fertil Steril. 1984 Jul;42(1):34–8.

53. Frydman R, Rainhorn J D, Forman R, Belaisch-Allart J, Fernandez H, Lassalle B, Testart J. Programmed oocyte retrieval during routine laparoscopy and embryo cryopreservation for later transfer Am J Obstet Gynecol. 1986 Jul;155(1):112–7. doi: 10.1016/0002-9378(86)90091-8.

5. Frydman R, Forman R, Rainhorn J D, Belaisch-Allart J, Hazout A, Testart J. A new approach to follicular stimulation for in vitro fertilization: programed oocyte retrieval Fertil Steril. 1986 Oct;46(4):657–62. doi: 10.1016/s0015-0282(16)49644-5.

54. Gizzo S, Andrisani A, Noventa M, Quaranta M, Esposito F, Armanini D, et al. Menstrual Cycle Length: A Surrogate Measure of Reproductive Health Capable of Improving the Accuracy of Biochemical/Sonographical Ovarian Reserve Test in Estimating the Reproductive Chances of Women Referred to ART. Reprod Biol Endocrinol (2015) 13:28. doi: 10.1186/s12958-015-0024

55. Younis JS, Iskander R, Fauser BCJM, Izhaki I. Does an Association Exist Between Menstrual Cycle Length Within the Normal Range and Ovarian Reserve Biomarkers During the Reproductive Years? A Systematic Review and Meta-Analysis. Hum Reprod Update (2020) 26:904–28. doi: 10.1093/humupd/dmaa013

56. Oehninger S, Nelson SM, Verweij P, Stegmann BJ. Predictive Factors for Ovarian Response in a Corifollitropin Alfa/GnRH Antagonist Protocol for Controlled Ovarian Stimulation in IVF/ICSI Cycles. Reprod Biol Endocrinol (2015) 13:117. doi: 10.1186/s12958-015-0113-1.

57. Xinyang Zhao, Xu Zhang, Shanshan Wu, Jichun Tan. Association Between the Ratio of Ovarian Stimulation Duration to Original Follicular Phase Length and In Vitro Fertilization Outcomes: A Novel Index to Optimise Clinical Trigger Time Front Endocrinol (Lausanne). 2022 Jul 25;13:862500. doi: 10.3389/fendo.2022.862500. eCollection 2022.

